# Transmission of SARS-CoV-2 on aircraft: A scoping review

**DOI:** 10.1101/2024.10.22.24315911

**Authors:** Constantine I. Vardavas, Katerina Nikitara, Katerina Aslanoglou, Apostolos Kamekis, Nithya Ramesh, Emmanouil Symvoulakis, Revati Phalkey, Jo Leonardi-Bee, Varvara Mouchtouri, Christos Hadjichristodoulou, Agoritsa Baka, Favelle Lamb, Jonathan E. Suk, Emmanuel Robesyn

## Abstract

**Introduction:** The assessment of empirical epidemiological data is needed to assess the transmissibility of SARS-CoV-2 in aircraft settings. This review summarises reported contact-tracing data and evaluates the secondary attack rates (SAR) and factors associated with SARS-CoV-2 transmission in aircraft, to provide insight for future decision making in the context of future respiratory pandemics.

**Methods:** This scoping literature review assessed studies published between December 2020 to November 2023 in Ovid Medline, Embase and Cochrane Library databases. The inclusion criteria were based on the PCC framework (P-Population, C-Concept, C-Context). The study population was restricted to passengers and crew (population) to assess transmission (concept) in an aircraft setting (context).

**Results:** Thirty-one studies which assess SARS-CoV-2 transmission in 521 domestic and international flights were included in this systematic review. The SAR reported in the studies with an identified index case ranged from 0% to 16%. Significant variation in the reporting across studies was noted. Overall, the studies reported that using face masks or respirators by passengers and crew members during flight seemed to be a possible strategy for mitigating SARS-CoV-2 transmission while sitting within close proximity to index cases (≤2 seats in every direction) was associated with a higher SAR.

**Conclusions:** Our results are consistent with sporadic clusters happening onboard aircraft. Close proximity to COVID-19 cases within the aircraft was associated with a higher SAR. Our findings further underscore the need for a systematic approach to examining and reporting SARS-CoV-2 transmission onboard aircraft. This evidence may assist policymakers and transportation authorities in the development of emergency preparedness measures and travel guidance during the post-pandemic COVID-19 era.

## INTRODUCTION

Aerosol and droplet transmission and contact with infected surfaces may all contribute to the onboard transmission of SARS-CoV-2, as in other closed environments [1]. Additionally, congregation during flights as well as during boarding and disembarking may also affect the likelihood of infection transmission within aircraft settings [2]. Measures such as mask use, screening, restriction of in-flight services, physical distancing measures, along with the use of high ventilation rates and high-efficiency particulate air (HEPA) filtration in the aircraft have been actions implemented by the aviation industry aiming to mitigate transmission within aircraft settings [3].

To better understand aircraft transmission, studies have been developed based on the dynamics of other respiratory infectious diseases such as influenza, and SARS-CoV-1, or have used experimental aerosol dispersion or modelling data to estimate the possibility and likelihood of in-flight transmission [4, 5]. While previous systematic reviews using studies published up to 2022 indicated that SARS-CoV-2 could be transmitted during aircraft travel [6, 7], we sought further epidemiological data for assessing the transmissibility of SARS-CoV-2 in aircraft, taking the entire duration of the COVID-19 pandemic and the most recent evidence into account. This review summarises reported contact-tracing information and provides and overview of reported secondary attack rates (SAR) and factors associated with in-flight SARS-CoV-2 transmission, that could be used to support both study design and public health decision-making within the context of future respiratory pandemics.

## METHODS

The scoping review was conducted according to the PRISMA (Preferred Reporting Items for Systematic Reviews and Meta-Analysis) [8] and MOOSE (Meta-analyses Of Observational Studies in Epidemiology) guidelines [9]. The protocol was not pre-registered in a database for systematic reviews.

### Outcomes and inclusion/exclusion criteria

Peer-reviewed studies published between December 1, 2020 and November 3, 2023 were identified through systematic electronic searches using OVID Medline, EMBASE, and the Cochrane Library. While a repository of reviews and not original studies, the Cochrane Library was searched so as to assess the reference lists of potential relevant reviews systematic and non-systematic literature reviews. The detailed search strategies are presented in **Supplementary - Appendix 1**. Predefined inclusion and exclusion criteria were used to determine the eligibility of the studies based on the PCC framework (P-Population, C-Concept, C-Context). The study population was restricted to passengers and crew members of the aircraft (population) to assess transmission (concept) in an aircraft setting (context). The exposure period was defined to include, apart from the flight duration, also pre-boarding and post-boarding time at the airport and time in transit (jointly hereon referred to as ‘flight-associated’). The primary outcome of our review was the existence of transmission of SARS-CoV-2, estimated through the Secondary Attack Rate (SAR) or other indexes of secondary transmission. An event of flight-associated transmission was defined as SARS-CoV-2 infection with no other reported source of infection. The classifications of index cases, secondary cases and the methods for case definition and diagnosis were extracted from the original studies. Modelling studies, experimental lab studies and opinion pieces were excluded from the review.

### Study selection

Studies identified from the searches were uploaded into a bibliographic database and duplicates were removed. Initially, a pilot training title/abstract screening process was used, where a random sample of 100 titles was independently screened for eligibility by two reviewers to enable consistency in screening and identify areas for amendments in the inclusion criteria. A high measure of inter-rater agreement was achieved (percentage agreement >90%), and hence the remaining titles were distributed between the two reviewers. For the full-text screening, a similar process was followed. Ten randomly selected studies were independently screened for eligibility by two reviewers, showing high agreement, after which the remaining of the full texts were subsequently distributed between the two reviewers for separate screening. Any disagreements were independently assessed by a third reviewer.

### Data extraction, synthesis, and presentation

Data extracted were related to the study design, study setting flight characteristics (domestic, international), sample characteristics (crew, passengers), number of cases/contacts traced/tested, definition of index cases and contacts, the variant of SARS-CoV-2, NPIs implemented before, during or following air travel, and numerical outcome measures including SAR also other statistical indexes of secondary transmission noted in the original manuscripts such as risk ratios (RR) and odds ratios (OR).

### Assessment of Risk of Bias

The nine point risk of bias tool developed by Leitmeyer & Adlhoch [10] was used to evaluate the risk of bias of the included studies, with the quality was categorised as low, medium or high based on the individual score of each study (0–3 low, 4–6 medium, 7–9 high). Low quality studies were not removed from the assessment, but subsequently flagged as LQ for reader’s information. It is important to note that this tool was developed for the assessment of single flight studies and hence may not be directly applicable to those studies which report multiple flights. The appraisal was performed independently by two reviewers, and disagreements were assessed and resolved by a third reviewer.

## RESULTS

A total number of 3,169 studies were identified according to the specified selection criteria from Medline, Embase and the Cohrane Library for systematic reviews. After the removal of duplicates, 2,543 were screened against their title and abstract, and 125 studies were assessed for full-text eligibility. Through the assessment of complete texts, 31 studies were included in the final study list, 26 identified through systematic electronic searches using OVID Medline, EMBASE, and the Cochrane Library and 5 identified through the reference lists of the included articles [11–41]. The flowchart of the study selection is presented in **Figure 1**. Real-time polymerase chain reaction (RT-PCR) to identify COVID-19 cases and contacts was conducted in all studies. Flight duration, origin and destination, index cases, contacts traced, data for secondary cases and SAR were available for most studies and are summarised in **Tables 1, 2 & 3**. The risk of bias assessment of the included studies are presented in detail in **Supplementary – Appendix 2**. All studies were of moderate or high quality, with the exception of one single flight study [29] and three multiple flight studies [17, 33, 41].

**Table 1.** Methodological and epidemiological investigation characteristics of included studies (n=31) assessing flight-associated transmission of SARS-CoV-2 published up to November 2023.

| AUTHOR/YEAR | DIAGNOSIS OF THE PRIMARY OUTCOME | TIMEFRAME/FOLLOW UP | CONTACT TRACING PROCESS & NPIs FOR CASE MANAGEMENT AND PREVENTION | FLIGHT RELATED NPIs |
| --- | --- | --- | --- | --- |
| Dhanasekaran et al., 2021 [11] | RT-PCR + genome sequencing | April 2021/ 30 days post-arrival | All passengers underwent a PCR test upon arrival<br>All passengers were obligated to quarantine for 21 days upon the arrival and were routinely tested. For passengers sharing accommodations with positive cases, quarantine periods were extended to 30 days of post-arrival. | **Predeparture**<br>Health declaration and a negative nucleic acid test within 72 hours of departure. Thermal screening and social distancing during check-in and boarding.<br>**In-Flight**<br>Mandatory mask use. Airline staff members were equipped with gowns, gloves, face masks, and face shields. The flight was at 85% capacity.<br>**Upon Arrival**<br>All passengers were tested by RT-PCR. |
| Guo et al., 2022 [12] | RT-PCR | June1 – August 1 2020 | All passengers underwent a PCR test upon arrival and were medically observed during the 14 day quarantine. | **Predeparture**<br>Facemasks were not mandatory<br>**In-flight**<br>Facemasks was not mandatory. The flight was at 75% occupancy in business, 100% in premium economy and 67% in economy.<br>**Upon Arrival**<br>All passengers were screened for body temperature by thermal imaging and were required to declare any COVID-19 symptoms. |
| Hu et al., 2021 [13] | RT-PCR or Rapid tests | January 4 – March 14 2020 | Close contacts (passenger who seated within three rows to the confirmed cases) were traced. | Data on usage of personal protective equipment such as face masks and goggles during the flight were not available. |
| Khahn et al., 2020 [14] | RT-PCR | March 1, 2020/ 13 days post arrival | All passengers who were followed up underwent a PCR test during contact tracing.<br>All cases, and their close contacts, were required to quarantine for 14 days and checked twice daily for clinical symptoms/signs | **Predeparture**<br>Facemasks were not mandatory<br>**In-flight**<br>Facemasks was not mandatory. The flight was at 75% occupancy in business, 100% in premium economy and 67% in economy.<br>**Upon Arrival**<br>All passengers were screened for body temperature by thermal imaging and were required to declare any COVID-19 symptoms. |
| Moek et al., 2022 [17] | RT-PCR | Phase 1: January 23, 2020- March 17 2020<br>Phase 2: June 5 -August 10, 2020 | Close contacts (passenger who seated within two rows to the confirmed cases) were traced. | **Pre departure**<br>Phase 1: Non mandatory masks<br>Phase 2: Mandatory masks<br>**In flight**<br>Phase 1: Non mandatory masks<br>Phase 2: Mandatory masks |
| Williamson et al., 2022 [20] | RT-PCR | June 26 2021 / 14 days post arrival | Isolation and contact tracing of all cases and 14-day quarantine and serial SARS-CoV-2 PCR testing of all close contacts. Additional questionnaire data were collected. | **Predeparture**<br>Mandatory face mask use<br>**In-Flight**<br>Mandatory face mask use. The flight was at 87.5% occupancy in economy and 78.6% in business |
| Zhang et al., 2021 [21] | RT-PCR | March 2020/ until August 1 2020 | All passengers were quarantined for 14 days. Suspected cases underwent a PCR test, while all other passengers were medically observed during the 14-day quarantine. Symptomatic passengers were transferred to a designated temporary place of isolation, while suspected cases were transferred to the designated hospitals where RT-PCR, chest radiograph, and routine blood tests were performed. Families were staying together. | **Predeparture**<br>Health declaration<br>**In-Flight**<br>Non-mandatory mask use. Almost every passenger used a face mask, and some wore medical protective clothing and goggles.<br>**Upon Arrival**<br>Health declaration and health inspection, including body temperature screening. |
| Bae et al., 2020 [15] | RT-PCR | March 31 2020/ until April 15 2020 | All passengers were quarantined for 14 days at a government quarantine facility completely isolated from one another and underwent routinely medical examination. All passengers underwent a PCR test on day 1 and day 14 of quarantine and were medically observed twice per day during quarantine. | **Predeparture**<br>Medical staff performed physical examinations, medical interviews, and body temperature checks outside the airport before boarding. Passengers were kept 2 m apart during pre-boarding.<br>**In-Flight**<br>Mandatory face mask use. N95 respirators were provided, which most passengers wore except at mealtimes and when using the toilet. |
| Eldin et al., 2020 [16] | RT-PCR | March 2020/ No follow up | Contact investigation by telephone | Unclear |
| Murphy et al., 2020 [18] | RT-PCR + genome sequencing | Summer 2020/ Outbreak was declared over 28 days after the last date of symptom onset. | Close contacts (seated within two seats in every direction from the first cases notified) and cabin crew were initially traced and when possible underwent a PCR test. The remaining passengers were offered testing where contactable. | **In-Flight**<br>Non-mandatory mask use. A mask was worn during the flight by nine flight cases, not worn by one (a child), and unknown for three. The airplane was at 17% occupancy. |
| Quach et al., 2021 [19] | RT-PCR | March 1 2020/ until March 16 2020 | Contact tracing started four days after the arrival of the flight. Primary contacts (all co-passengers and crew members) were traced and those who could be reached were interviewed. All passengers who had transited out of Vietnam were contacted through border health control authorities. | **In-Flight**<br>Non-mandatory mask use.<br>**Upon Arrival**<br>Health declaration and body temperature screening. |
| Blomquist et al., 2021 [23] | RT-PCR + genome sequencing | January – March 12 2020 | Identification of cases with recent flight history and their contacts. Travellers from high-incidence areas were isolated, and, where reachable, contacts were put under passive surveillance for 14 days from the day of the flight. If they developed symptoms, they were asked to inform public health services, and get tested. | **In-Flight**<br>Non-mandatory mask use. |
| Yang et al., 2020 [26] | RT-PCR | January 24 2020/<br>14 days from the day of<br>the flight | All passengers and crew members were isolated/quarantined and underwent routine medical cheques at hotels near the airport for 14 days. | **Predeparture**<br>Strict pre-flight screening of fever or respiratory symptoms.<br>**In-Flight**<br>Non-mandatory mask use. Most passengers did not take any precautionary measures against possible exposure to SARS-CoV-2 during the flight, while all flight attendants wore masks. The index case was not wearing a face mask.<br>**Upon Arrival**<br>Screening for fever and respiratory symptoms in passengers and crew members. |
| Butt et al., 2021 [24] | RT-PCR | August 5 2020 - February<br>22 2021 | All passengers were tested upon arrival and medically observed during the 14-day quarantine. Repeat testing was performed 1 week after arrival and those who tested negative were released from quarantine. Those with a positive test were immediately transferred to a designated isolation facility or a hospital. | **Predeparture**<br>Medical screening for symptoms<br>**In-Flight**<br>Mandatory mask use, frequent disinfection of the aircraft, encouraging use of hand sanitizers.<br>**Upon Arrival**<br>Medical evaluation at the airport and RT-PCR |
| Nir Paz et al., 2020 [25] | RT-PCR | February 20 2020/14 days | All passengers were tested upon arrival and medically observed during the 14-day quarantine (6 tests while in quarantine). | **Predeparture**<br>At least 1 negative RT-PCR.<br>**In-Flight**<br>Mandatory mask use. Medical masks and respirator (FFP2) wearing which were replaced every 3 hours for the duration of the flight. Masks were taken off only for eating and drinking.<br>**Upon Arrival**<br>All passengers were tested. |
| Schwartz et al., 2020 [31] | RT-PCR | January 22 2020 | The public was notified through the media that the index case was symptomatic during the flight. Passengers and crew members who were not from Ontario were referred to their home jurisdictions for follow-up. Close contacts (individuals sitting within 2 m of the index case) received active daily contact monitoring by local public health officials for 14 days from the flight's arrival in Toronto. Non-close-contact passengers were advised to self-monitor and contact public health if they became symptomatic. | **In-Flight**<br>Mandatory mask use. |
| Choi et al., 2020 [28] | RT-PCR + genome<br>sequencing | March 9 2020/<br>up to 11 days after arrival | Publicly available data – only symptomatic identified. Airport screening and quarantine had not been implemented in Hong Kong yet. | Unclear |
| Hoehl et al., 2020 [29] | RT-PCR | March 9 2020/<br>6-9 weeks | Passengers were interviewed by phone and PCR tests 4 to 5 weeks after the flight. A semiquantitative SARS-CoV-2 IgG antibody test was offered to all passengers who had been seated within 2 rows of the index cases and to those who reported to have been symptomatic. | **In-Flight**<br>No measures to prevent transmission (e.g., wearing of masks) had been applied. |
| Kong et al., 2021 [30] | RT-PCR | 16-28 January 2020/<br>14 days | Epidemiological investigation was conducted through the extraction of travel and clinical information. 14-day quarantine for suspected cases and isolation for confirmed cases. | Unclear |
| Swadi et al., 2021 [32] | RT-PCR + genome sequencing | September 28 2020/<br>14 days | All passengers were tested and medically observed during the 14-day quarantine. Managed isolation and quarantine at the border. PCR diagnostic testing for SARS-CoV-2 on day 3 and again on day 12 if the previous test result was negative. | **In-Flight**<br>Non-mandatory mask use. Some passengers wore gloves. |
| Chen et al., 2020 [27] | RT-PCR | February 15 2020/14 days | All passengers were tested and medically observed during the 14-day quarantine. All passengers were regarded as close contacts and required to follow medical isolation and observation protocols for at least 14 days. | **In-Flight**<br>The aircraft was at 89% occupancy. All passengers were required to take their temperature before deplaning. |
| Eichler et al., 2021 [34] | RT-PCR + genome sequencing | August 27 2020 | All passengers were tested and medically observed during the mandatory 14-day hotel managed isolation and quarantine for all passengers. | **Predeparture**<br>Testing for SARS-CoV-2 was not mandatory.<br>**In-Flight**<br>Mandatory mask use. The aircraft was at ≈35% occupancy. Passengers were evenly spaced throughout the aircraft. |
| Lv et al., 2021 [35] | RT-PCR | 9-10 June 2021/14 days | All passengers were tested and medically observed during the mandatory 14-day quarantine. | **Predeparture**<br>Negative PCR and IgM assays done at any two designated testing institutions (Pathcare, AMPATH, Lancet) within 48 h before boarding.<br>**In-Flight**<br>Mandatory mask use. |
| Pavli et al., 2020 [36] | RT-PCR | February 26 2020 - March 9 2020 | Contact tracing of close contacts (distance of <2 m for >15 min), including passengers seated two seats in all directions around the index case and all crew members and persons who had close contact with the index case (e.g., travel companions or persons. Advice was provided to all close contacts for 14-day self-quarantine following their last exposure and self-monitoring for respiratory symptoms and/or fever, providing care). | Unclear |
| Speake et al., 2020 [37] | RT-PCR + genome sequencing | March 19 2020/14 days | All close contacts were informed of their potential exposure to SARS-CoV-2 and directed to quarantine themselves for 14 days. Attempts were made to notify all remaining passengers of their potential exposure. Persons experiencing symptoms were tested with PCR. | **In-Flight**<br>Non-mandatory mask use. |
| Toyokawa et al., 2021 [38] | RT-PCR | March 23 2020/ April 26–29, 2020, 34 days after the index case was reported | Primarily contact tracing of passengers who sat within two rows from the index patient. The contact tracing was then expanded to more passengers. Passengers were notified by telephone about their potential exposure to SARS-CoV-2 infection and were asked to self-quarantine and self-monitor until 14 days after the flight. | **In-Flight**<br>Non-mandatory mask use. The aircraft was at 80.2% occupancy. |
| Draper et al., 2020 [33] | RT-PCR | March 1 - April 30 2020 | Rapid and thorough contact tracing of close contacts. Close contacts were quarantined for 14 days and undergone active daily monitoring for symptoms. | Unclear |
| Park et al., 2020 [40] | RT-PCR | February 8 2020 | Contact tracing of COVID-19 cases was conducted from 1 day before symptom onset or 1 day before the case was sampled. There was a limitation of contact tracing foreign passengers. Confirmed COVID-19 cases were admitted to 4 designated hospitals. Pilgrim travellers who tested negative were home quarantined for 14 days. Contacts who experienced symptoms within 14 days of home quarantine were tested for SARS-CoV-2. | **In-Flight**<br>Non-mandatory mask use. |
| White et al., 2022 [41] | RT-PCR | December 2020 | Close contacts of the primary cases were identified and tested with PCR. Where close contacts had no SARS-CoV-2 swab result, the Covid Care Tracker was used for information. PCR testing was offered to incoming travellers from 'red' regions and from the United Kingdom on day 5 after arrival. | **Predeparture**<br>Negative pre-departure PCR test within the 72 h before arrival in Ireland was required of passengers from 'orange' regions within the EU. |
| Zhang XA et al., 2021 [22] | RT-PCR + genome sequencing | 22-24 January 2020/ 14 days | Close contacts were separately quarantined and monitored for symptoms. Symptomatic contacts were tested with PCR. | Unclear |
| Ngeh et al., 2022 [39] | RT-PCR + genome sequencing | 1 July 2020 / 14 days | Passengers were quarantined and tested for 14 days after the flight.<br>All passengers were isolated in quarantine hotels and monitored for symptoms. PCR was performed on Day 2 and Day 12 after arrival and if symptoms were noted. | **Predeparture**<br>Mandatory mask use, social distancing<br>**In-Flight**<br>Mandatory mask use. Travel packs, including gloves, facemasks, hand sanitizer and cleaning maks were provided by the airline. Crew wore masks, eye goggles, gloves and protective gowns.<br>**Upon Arrival**<br>All passengers were isolated in quarantine hotels and were tested with PCR. |

**Table 2.** Single flight details and Secondary Attack Rates (SARs) of flight-associated SARS-CoV-2 transmission, grouped by risk of Bias assessment, published up to November 2023.

| AUTHOR/YEAR | NUMBER OF FLIGHTS | FLIGHT DURATION | ORIGIN | DESTINATION | INDEX CASES | CONTACTS TRACED | SECONDARY CASES | Indexes of secondary transmission |
| --- | --- | --- | --- | --- | --- | --- | --- | --- |
| High Quality |  |  |  |  |  |  |  |  |
| Dhanasekaran et al., 2021 [11] | Single (n=1) | 6 h | New Delhi, India | Hong Kong | Total: At least 7 (Kappa: 3, Delta: 1, Alpha: 3) | 43 sequenced | Total: 36 | Unclear |
| Ngeh et al., (53) | Single (n=1) | 10h | Dubai, United Arab Emirates | Perth, Australia | Total: 2 | 90 pax | Total: 15 | SAR of 16% (15/93)<br>RR of 2.79, 95%CI: 1.09-7.15 for not a mask at all times<br>RR 2.71, 95%CI: 1.11-6.61 for sitting <2 seats from the index cases, RR 6.21, 95%CI: 1.71-22.24 for sitting <2 seats from the index case and not wearing a mask at all times<br>OR 7.16, 95%CI: 1.66-30.85 after controlling for mask use, seat proximity and time spent at the airport |
| Bae et al., 2020 [15] | Single (n=1) | 11 h | Milan, Italy | South Korea | Total: 6<br>Symptomatic: 0<br>Asymptomatic: 6<br>Pre-symptomatic: 0 | 293 pax + 18 crew | Total: 1<br>Symptomatic: 1<br>Asymptomatic: 0 | 1/311 (0.32%) |
| Quach et al., 2021 [19] | Single (n=1) | 10 h | London, UK | Hanoi, Vietnam | Total: 1<br>Symptomatic: 1<br>Asymptomatic: 0<br>Pre-symptomatic: 0 | 168 pax + 16 crew | Total: 14 px + 1 crew | 15/184 (8.2%) |
| Yang et al., 2020 [26] | Single (n=1) | 5 h | Singapore | Zhejiang | Total: 1<br>Symptomatic: 1<br>Asymptomatic: 0<br>Pre-symptomatic: 0 | 324 pax + crew | Total: 9 (+2 not interviewed)<br>Symptomatic: 7<br>Asymptomatic: 2 | 9 (+2 probably)/324 (2.8%) |
| Butt et al., 2021 [24] | Unclear | Not reported | International | Qatar | Total: 3789 | 348,385 | Total: 4447 | 4447/348,384 (1.3%) |
| Chen et al., 2020 [27] | Single (n=1) | Not reported | Singapore | Hangzhou, China | Total: 15<br>Symptomatic: 9<br>Asymptomatic: 6<br>Pre-symptomatic: 0 | 331 | Total: 1<br>Symptomatic: 1<br>Asymptomatic: 0 | 1/331 (0.30%) |
| Lv et al., 2021 [35] | Single (n=1) | Not reported | South Africa | Shenzhen, China | **Total:** 1 (Delta variant) (+ 5 primary cases)<br>**Symptomatic:** 0<br>**Asymptomatic:** 0<br>**Pre-symptomatic:** 1 | 197 | **Total:** 20 | 20/197 (10.2%) |
| Khahn et al., 2020 [14] | Single (n=1) | 10 h | London | Hanoi, Vietnam | **Total:** 1 | 201 pax + 16 crew | **Total:** 15 (14 pax + 1 crew) | SAR of 62% (13/21) within business class passengers<br>SAR of 7.4% overall (16/217) |
| Medium Quality |  |  |  |  |  |  |  |  |
| Williamson et al., 2022 [20] | Single (n=1) | 81 min | Gold Coast, Australia | Sydney, Australia | **Total:** 1 | 139 pax + 6 crew | **Total:** 11 | 6.9% (10/145) |
| Murphy et al., 2020 [18] | Single (n=1) | 7.5 h | International | Ireland | **Total:** 1 to 7 | 37 | **Total:** 4 to 12 | Unclear<br>(Unknown source cases with scenarios ranging between 4/41 (9.8%) to 12/48 (25%) if no cases were within the incubation period *) |
| Eldin et al., 2020 [16] | Single (n=1) | Not reported | Bangui, Africa | Paris, France | **Total:** 1<br>**Symptomatic:** 1<br>**Asymptomatic:** 0<br>**Pre-symptomatic:** 0 | 3 | **Total:** 1<br>**Symptomatic:** 1<br>**Asymptomatic:** 0 | Unclear |
| Nir Paz et al., 2020 [25] | Single (n=1) | 13.5 h | Japan | Israel | **Total:** 2<br>**Symptomatic:** 0<br>**Asymptomatic:** 2<br>**Pre-symptomatic:** 0 | 9 pax + 4 crew | **Total:** 0 | 0 |
| Schwartz et al., 2020 [31] | Single (n=1) | 15 h | Wuhan, China | Toronto (via Guangzhou) | **Total:** 2<br>**Symptomatic:** 2<br>**Asymptomatic:** 0<br>**Pre-symptomatic:** 0 | 26 (Close contacts sitting within 2 m of the index case + crew members) | **Total:** 0 | 0 |
| Choi et al., 2020 [28] | Single (n=1) | ≈15 h | Boston, USA | Hong Kong, China | **Total:** 2<br>**Symptomatic:** 2<br>**Asymptomatic:** 0<br>**Pre-symptomatic:** 0 | 2 crew (only symptomatic COVID-19 cases and contacts were traced) | **Total:** 2 crew<br>**Symptomatic:** 1<br>**Asymptomatic:** 1 | Unclear |
| Swadi et al., 2021 [32] | Single (n=1) | 18 h | Dubai | New Zealand via Kuala Lumpur (no passengers entered or exited) | **Total:** 2<br>**Symptomatic:** 2<br>**Asymptomatic:** 0<br>**Pre-symptomatic:** 0 | 84 pax | **Total:** 4 (1 probably got infected during quarantine)<br>**Symptomatic:** 2<br>**Asymptomatic:** 2 | 4/84 (4.76%) |
| Eichler et al., 2021 [34] | Single (n=1) | ≈18 hour | Delhi, India | Christchurch, New Zealand via Nadi, Fiji | **Total:** 1-2<br>**Symptomatic:** 1<br>**Asymptomatic:** 1<br>**Pre-symptomatic:** 0 | Unknown | **Total:** 1 to 2<br>**Symptomatic:** 2<br>**Asymptomatic:** 0 | Unclear |
| Eichler et al., 2021 [34] | Single (n=1) | Not reported | Christchurch | Auckland | **Total:** 1<br>**Symptomatic:** 0<br>**Asymptomatic:** 1<br>**Pre-symptomatic:** 0 | Unknown | **Total:** 1<br>**Symptomatic:** 1<br>**Asymptomatic:** 0 | Unclear |
| Speake et al., 2020 [37] | Single (n=1) | 5 h | Sydney, Australia | Perth, Australia | **Total:** 18 (11 possibly infectious in flight) | 213 | 8 + 2 possible | 8/213 (3.8%) |
| Toyokawa et al., 2021 [38] | Single (n=1) | 2 h | City in Japan | Okinawa, Japan | **Total:** 1 | 145 | **Total:** 14 confirmed + 6 probable | 9.7%. If probable cases were included, the attack rate would be 13.8% |
| Park et al., 2020 [40] | Single (n=1) | Not reported | Israel | Korea | **Total:** 2 (+ 18 primary cases) | Unknown | **Total:** 2 | Unclear |
| Low Quality |  |  |  |  |  |  |  |  |
| Hoehl et al., 2020 [29] | Single (n=1) | 4 h and 40 m | Tel Aviv, Israel | Frankfurt, Germany | **Total:** 7<br>**Symptomatic:** 4<br>**Asymptomatic:** 1<br>**Pre-symptomatic:** 2 | 71 | **Total:** 2 (these transmissions may have also occurred before or after the flight)<br>**Symptomatic:** 1<br>**Asymptomatic:** 1 | 2/71 (2.8%) |

### Secondary Attack Rate (SAR) of SARS-CoV-2 in aircraft

Among the 31 studies included in this systematic review, 521 domestic and international flights were examined for SARS-CoV-2 transmission **(Table 2 & 3), with the results grouped based on study quality;** with ten studies reporting on multiple flights **(Table 3)**. The search results included flight-associated outbreaks occurring from 24 January 2020 to the 26^th^ June 2021.

**Table 3.**
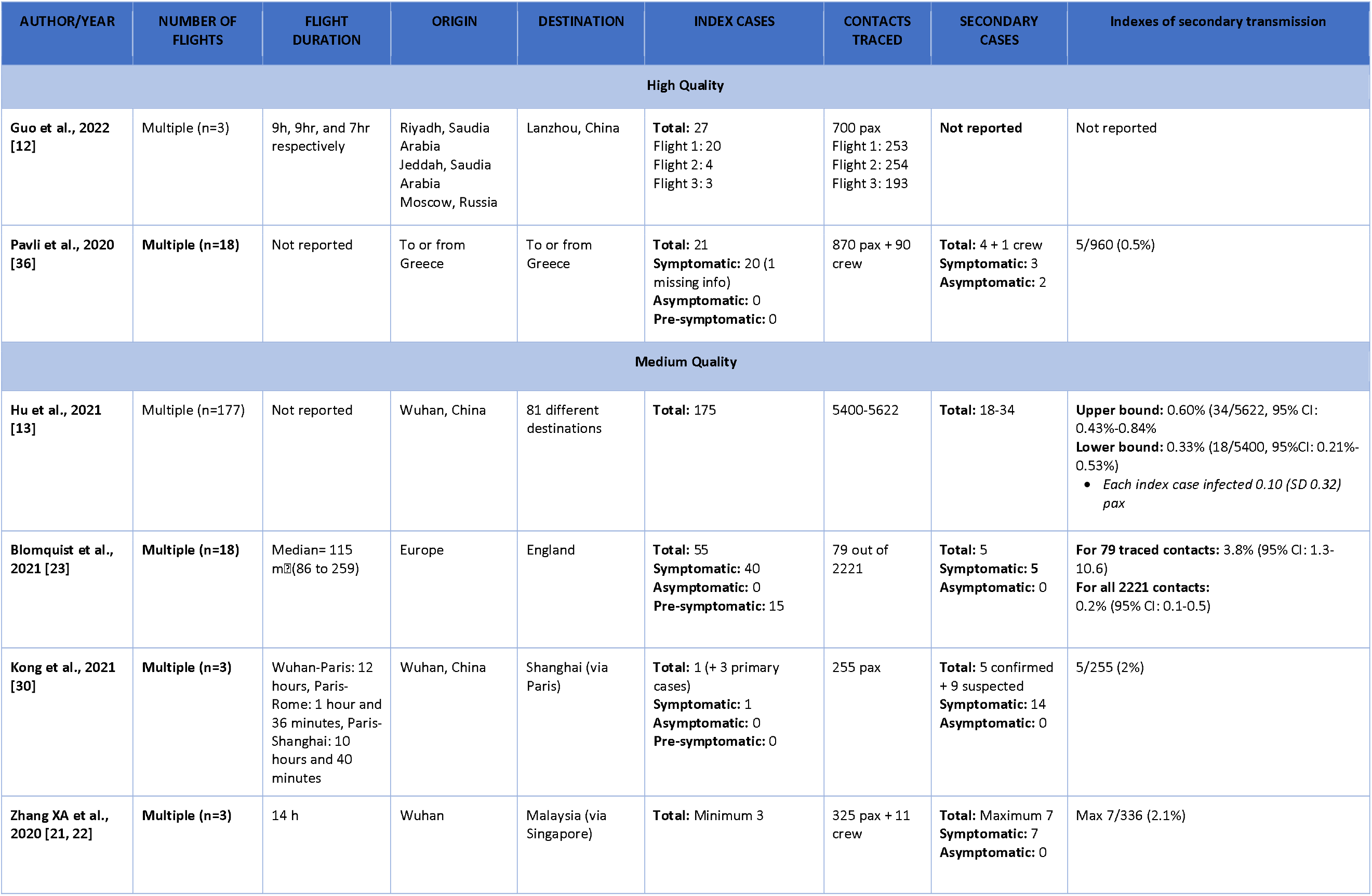
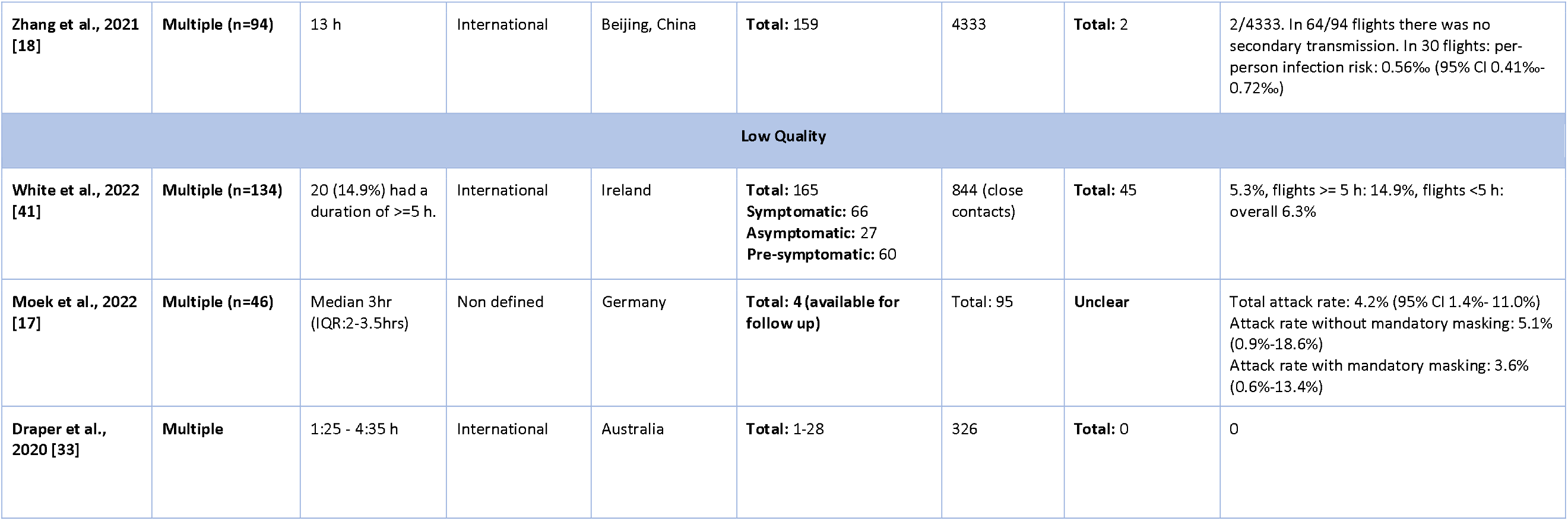
Multiple flight details and Secondary Attack Rates (SARs) of flight-associated SARS-CoV-2 transmission, grouped by risk of bias assessment, published up to November 2023.

| AUTHOR/YEAR | NUMBER OF FLIGHTS | FLIGHT DURATION | ORIGIN | DESTINATION | INDEX CASES | CONTACTS TRACED | SECONDARY CASES | Indexes of secondary transmission |
| --- | --- | --- | --- | --- | --- | --- | --- | --- |
| High Quality |  |  |  |  |  |  |  |  |
| Guo et al., 2022 [12] | Multiple (n=3) | 9h, 9hr, and 7hr respectively | Riyadh, Saudia Arabia<br>Jeddah, Saudia Arabia<br>Moscow, Russia | Lanzhou, China | Total: 27<br>Flight 1: 20<br>Flight 2: 4<br>Flight 3: 3 | 700 pax<br>Flight 1: 253<br>Flight 2: 254<br>Flight 3: 193 | Not reported | Not reported |
| Pavli et al., 2020 [36] | Multiple (n=18) | Not reported | To or from Greece | To or from Greece | Total: 21<br>Symptomatic: 20 (1 missing info)<br>Asymptomatic: 0<br>Pre-symptomatic: 0 | 870 pax + 90 crew | Total: 4 + 1 crew<br>Symptomatic: 3<br>Asymptomatic: 2 | 5/960 (0.5%) |
| Medium Quality |  |  |  |  |  |  |  |  |
| Hu et al., 2021 [13] | Multiple (n=177) | Not reported | Wuhan, China | 81 different destinations | Total: 175 | 5400-5622 | Total: 18-34 | Upper bound: 0.60% (34/5622, 95% CI: 0.43%-0.84%<br>Lower bound: 0.33% (18/5400, 95%CI: 0.21%-0.53%)<br>• Each index case infected 0.10 (SD 0.32) pax |
| Blomquist et al., 2021 [23] | Multiple (n=18) | Median= 115 min (86 to 259) | Europe | England | Total: 55<br>Symptomatic: 40<br>Asymptomatic: 0<br>Pre-symptomatic: 15 | 79 out of 2221 | Total: 5<br>Symptomatic: 5<br>Asymptomatic: 0 | For 79 traced contacts: 3.8% (95% CI: 1.3-10.6)<br>For all 2221 contacts: 0.2% (95% CI: 0.1-0.5) |
| Kong et al., 2021 [30] | Multiple (n=3) | Wuhan-Paris: 12 hours, Paris-Rome: 1 hour and 36 minutes, Paris-Shanghai: 10 hours and 40 minutes | Wuhan, China | Shanghai (via Paris) | Total: 1 (+ 3 primary cases)<br>Symptomatic: 1<br>Asymptomatic: 0<br>Pre-symptomatic: 0 | 255 pax | Total: 5 confirmed + 9 suspected<br>Symptomatic: 14<br>Asymptomatic: 0 | 5/255 (2%) |
| Zhang XA et al., 2020 [21, 22] | Multiple (n=3) | 14 h | Wuhan | Malaysia (via Singapore) | Total: Minimum 3 | 325 pax + 11 crew | Total: Maximum 7<br>Symptomatic: 7<br>Asymptomatic: 0 | Max 7/336 (2.1%) |
| Zhang et al., 2021 [18] | Multiple (n=94) | 13 h | International | Beijing, China | Total: 159 | 4333 | Total: 2 | 2/4333. In 64/94 flights there was no secondary transmission. In 30 flights: per-person infection risk: 0.56‰ (95% CI 0.41‰-0.72‰) |
| Low Quality |  |  |  |  |  |  |  |  |
| White et al., 2022 [41] | Multiple (n=134) | 20 (14.9%) had a duration of >=5 h. | International | Ireland | Total: 165<br>Symptomatic: 66<br>Asymptomatic: 27<br>Pre-symptomatic: 60 | 844 (close contacts) | Total: 45 | 5.3%, flights >= 5 h: 14.9%, flights <5 h: overall 6.3% |
| Moek et al., 2022 [17] | Multiple (n=46) | Median 3hr (IQR:2-3.5hrs) | Non defined | Germany | Total: 4 (available for follow up) | Total: 95 | Unclear | Total attack rate: 4.2% (95% CI 1.4%- 11.0%)<br>Attack rate without mandatory masking: 5.1% (0.9%-18.6%)<br>Attack rate with mandatory masking: 3.6% (0.6%-13.4%) |
| Draper et al., 2020 [33] | Multiple | 1:25 - 4:35 h | International | Australia | Total: 1-28 | 326 | Total: 0 | 0 |

The extensive heterogeneity in definitions precluded the ability to perform pooled analyses and hence the results on SAR are presented below narratively. Within the current review, in four studies, the highest number of index cases were identified in studies looking at single flights rather than those providing evidence from multiple flights. These studies included two domestic flights reviewed by Speake et al. [MQ] [37] and Chen et al. [HQ] [27] and two international flights by Dhanasekaran et al. [11] and Hoehl et al. [29]. Regarding the domestic flights, Speake et al., conducted a cohort study on a five-hour domestic flight from Sydney to Perth, Australia, with a total of 241 passengers and 18 index cases, of which 11 had been infectious during the flight [37]. Chen et al. also conducted a cohort study on a flight from Singapore to Hangzhou, China, with 335 passengers and 11 crew members and reported 15 index cases, nine of which were symptomatic [27]. Regarding the studies looking at international flights, Dhanasekaran et al. [11] identified seven index cases in a large cluster of 59 cases that were linked to a single flight from New Delhi, India to Hong Kong, China, with 146 passengers. Finally, a case series conducted for a 4-hour flight from Tel Aviv, Israel to Frankfurt, Germany by Hoehl et al. (LQ) identified seven index cases (four symptomatic, two pre-symptomatic and one asymptomatic) [29]. All other studies referring to single flights reported a range of 1-3 index cases [17, 20, 22, 24, 26–28, 31, 33–35, 37, 39, 42, 43].

Of 31 studies included in the analysis, the reported SAR varied from 0% to 16%, three studies lacked the information needed to estimate flight-associated SAR [28, 34, 40], while one reported ranges of SAR estimates based on scenarios [18] and one based on travel class within the airplane [14]. Among the studies where SAR could be calculated, no secondary flight-associated transmission was reported in six studies [11, 16, 21, 25, 31, 33]. Eight flights noted SARs between 5-16.2%, all on single flights [35, 39] [14, 19, 32, 38, 41]. Finally, seven studies presented a SAR between 1-5% [17, 22, 24, 26, 29, 30, 36, 37] and six studies reported a SAR below 1% [13, 15, 21, 23, 27, 36]. Only four studies in our review provided explicit data considering the transmission of specific SARS-CoV-2 variants [11, 20, 35, 39], while studies performed early into the COVID-19 pandemic self-classified the strain as the initial wild-type SARS-CoV-2 variant [14, 17].

### Flight-related mitigation measures

Regarding NPIs implemented during the flights, mask use was stated to be mandatory in nine studies [11, 15, 20, 24, 25, 31, 34, 35, 39], while the use of masks was not obliged in 12 investigations [18, 21–24, 29, 31, 32, 36, 37, 39, 42], while one study performed a pre-post facemask regulation comparison of SAR [17]. Most of the included flights occurred in early 2020, therefore, this may in part explain this heterogeneity in the application of mask use. Of the studies reporting on the effectiveness of mitigation measures, Williamson et al., within the context of one flight reported that removal of mask during flight (P=0.03), including eating (RR5.3) and drinking (RR10.6) were associated with an increased risk of passengers becoming a case [20], while Ngeh et al., noted that passengers that did not wear a mask at all the time during the flight had a RR of 2.79, (95%C.I: 1.09-7.17) to be secondary case [39]. Among the first eight studies with the highest SARs [14, 18–20, 32, 35, 38, 39, 41], mask use was not mandatory in five of them [14, 18, 19, 32, 38], while the study by Moek et al., (LQ) noted that among fights after mandatory masking, the SAR among close contacts was 3.6% (95% CI 0.6%–13.4%) while 5.1% (95% CI 0.9%–18.6%) before masking [17].

Aircraft occupancy was described in seven studies [11, 14, 18, 20, 27, 34, 38], with four [10, 19, 20, 21] indicating more than 80% occupancy (range 17-89%). All passengers were tested upon arrival in three studies [11, 24, 25, 39], and screening for body temperature and respiratory symptoms was recorded in another four investigations [14, 19, 21, 26]. Passengers were quarantined for 14 to 21 days in 13 studies [13, 17–19, 24–26, 31–34, 41–43], while in one study [23] only travelers from high-incidence areas and their reachable contacts were isolated and quarantined and in six studies only close contacts or suspected cases were quarantined [30, 31, 33, 37, 38, 40].

### Distance from the index case

Ten studies reported data regarding the distance of secondary cases from the index cases in the aircraft [12, 14, 17, 19, 23, 29, 30, 32, 34, 36, 38, 39]. In general, sitting within two rows of the index case was used as a measure of proximity. Khahn et al noted that seating at a distance <2 seats away from the index case was associated with an RR of 7.3, 95%C.I: 1.2-46.2)[14], while Ngeh noted an RR of 2.7 for the same distance, which increased to 6.2 (95%C.I:1.74-22.24), among those that did not wear a mask at all times [39]. In the study of Blomquist et al. [23], from the five secondary cases, four were sitting within two rows of an index case and one five rows away. Similarly, Swadi et al. [32] reported four passengers as secondary cases, all seated within two rows of index cases. Also, in the study by Kong et al. [30], all nine confirmed and suspected secondary cases were seated within two rows from the index case. Among the 15 probable secondary cases described in the study of Quach et al. [19], 12 were in the travelling class as the index case, while 11 were sitting at a distance of ≤2 seats from the index case. Likewise, all secondary cases were sitting within two rows from the index case in the study of Hoehl et al. [29], Eichler et al. [34], and Pavli et al. [36]. Toyokawa et al. [38] stated that the passengers seated within two rows from the index patient was 4.8 (95% CI: 1.46–15.8) times more likely to get infected with SARS-CoV-2. Finally, in the study by Williamson et al., where the index case was a crew member, sitting where the index case predominately worked was associated with an increased risk of transmission [20]. In summary, for almost all studies presenting distance from the index cases, secondary cases were more likely to be within close proximity to the index case.

## DISCUSSION

During 2022, there was a significant rise in international tourism compared with the previous two years, with nearly 900 million international trips recorded worldwide, albeit still substantially lower than pre-pandemic levels [42, 43]. This review aimed to assess in-flight transmission of SARS-CoV-2, and indicated the existence of sporadic clusters of transmission, within which the measured SAR rate was found to range from 0% to 10.2%. Similar to our results from 521 reported flights, the findings of an earlier systematic review of Rosca et al., indicate that the SAR reported in published research (following up >80% of passengers and crew for 130 flights) varied between 0 and 8.2% [6], while a recent review by Lu et al., noted a SAR of 7.6%, ranging between 2.6% to 16.1%.

Previous research has suggested that mask-wearing may mitigate transmission [44, 45]. Within the context of our review and across the limited data that were available, when studies reported that most passengers and crew members used face masks during the flight, this strategy was deemed to be potentially effective in limiting the spread of SARS-CoV-2 [15, 21, 25, 31]. In contrast, within the studies when masks use was not implemented during the flight, the reported SAR was higher [19, 23, 37]. These results are further corroborated by recent modelling studies which indicate that FFP2/N95 mask use may decrease infection by 95-100%, while distancing measures – such as leaving the middle seat empty - may further reduce transmission [46]. Furthermore, although only one study in our review noted that facemasks were removed during food service [25], previous experimental work has indicated that the removal of masks, even for short periods on a long haul flight, such as for a 1-hour meal service, increases the average probability of infection by as much as 59%, compared to the situation where the mask is worn continuously [5].

Distance, based on studies of tracer gas/particle dispersion, is also considered a significant determinant of contaminant exposure on airplanes [47]. For example, results during the assessment of SARS-CoV-1 transmission in 2003 suggested that passengers within two rows of cases had a higher likelihood of infection compared to those seated further away [48]. According to our review, proximity to index cases was another significant parameter as passengers who were seated within two rows from the index cases were more likely to get infected. Our results also concur with the findings of Rosca et al., who noted that distance between the passengers and the index case on board could have impacted the spread of COVID-19 [6] and with the review of Lu et al., [7] that noted that the risk ratios of infection for passengers seated within and outside the two rows of the index cases were 5.64 (95% CI:1.94–16.40). While the role of HEPA filtration was not assessed within the context of this review as it is implemented by default in the aviation industry, it is possible that these filters may reduce generalized airborne transmission and hence limit direct transmission to close contacts [49].

With regards to flight duration, it is not clear from our review if it can affect the COVID-19 spread, as there were both long flights with secondary cases [31] and short flights (<5 h) with indications of transmission [38]. According to the systematic review of Rosca et al., who categorised flight duration as short, medium, or long, with a low or high proportion of secondary cases, transmission did not necessarily increase with flight duration [6]. More studies reported secondary cases of Kappa variant than of Alpha or Delta variants, although this is expected to depend mostly on the circulating variant at the time of the cluster. The infections were found despite RT-PCR tests being performed within 72 hours pre- departure, and personal protective measures were implemented during the flight [11]. In the study of Lv et al., the flight-associated transmission was from an index case who was infected by the Delta variant, which is known to have higher transmissibility compared to previous strains, with also in this study passengers required to have a 48-hour negative PCR test before departure, and mandatory mask use on board [35]. No conclusions can be drawn regarding transmissibility between variants based on the small number of reported observations of flight-associated transmission.

### Strengths and limitations

The systematic approach that was applied during study identification, data extraction, and risk of bias assessment are strengths of our study – including the recent November 2023 cut-off, which should allow for the identification of evidence collected for the duration of the COVID-19 pandemic. Despite these strengths, limitations of this review should be acknowledged. Firstly, there was no homogenous or systematic approach to the reporting of parameters that may be linked with flight-associated transmission. Secondly, we can not rule out publication bias as the reporting of larger clusters is to be expected, which if calculated, would decrease SARs. Thirdly, there was a broad handling of cases across studies where some of the studies did not screen asymptomatic index and secondary cases, while in others studies passengers were never traced successfully; therefore, the actual number of secondary cases remains unknown. Fourthly, there was significant heterogeneity in the in-parallel implemented NPIs across studies. Finally, as SARS-CoV-2 transmission was multifactorial, and potentially impacted by multiple factors such as passenger and crew individual behaviour and mobility, adherence to facemask use, vaccination status of participants, the transmissibility of the variants at the time of each study, and the proximity and duration of interactions between passengers, comparisons between studies cannot be directly made and the results should be interpreted with caution. Future studies should aim for a complete screening and evaluation of all passengers and crew members during follow-up, documenting aspects such as the genomics of the variants detected, distance from index cases, implemented pre and post-boarding procedures, and in-flight NPIs that are applied. The further standardisation of definitions and diagnostic procedures (including of index, primary, secondary, and close contact cases), across studies is further warranted. Despite the above limitations, this review provides a novel overview of the factors that may have impacted flight-associated transmission within the context of the COVID-19 pandemic, information which can be used in future emergency preparedness planning.

## CONCLUSIONS

Our results are consistent with sporadic clusters happening onboard aircraft. At the same time, close proximity to COVID-19 cases within the aircraft was associated with a higher SAR. Further research is still needed to better assess the impact of implementing NPIs during flight, boarding and disembarking on the transmission of SARS-CoV-2, including the type of masks used and ventilation. Our findings underscore the need for a systematic approach to examining and reporting SARS-CoV-2 transmission onboard aircrafts, which is required to decrease the variation in parameters reported between studies. This evidence may assist policymakers and transportation authorities in the development of emergency preparedness measures and travel guidance during the post-pandemic COVID-19 era.

## Data Availability

Data sharing is not applicable to this article as no new data were created or analysed in this study. 

## Acknowledgments

We would like to thank Katerina Papathanasaki, Stella Vogiatzidaki and Anastasia Manta for their assistance in data archiving.

## Funding

This report was produced under a service contract 17-ECD.12552, within Framework contract ECDC/2019/001 Lot 1B, with the European Centre for Disease Prevention and Control (ECDC), acting under the mandate from the European Commission. The information and views set out in this piece of work are those of the authors and do not necessarily reflect the official opinion of the Commission/Agency. The Commission/Agency do not guarantee the accuracy of the data included in this analysis. Neither the Commission/Agency nor any person acting on the Commission’s/Agency’s behalf may be held responsible for the use which may be made of the information contained therein.

**Conflicts of interest/Competing interests**

None to report.

## Supporting information

**S1 File. Search strategy for the assessment of flight-associated transmission of SARS-CoV-2.** (DOC)

**S2 File. Results of the Risk of Bias assessment of included studies related to flight associated transmission of SARS-CoV-2 as per the quality appraisal tool by Leitmeyer & Adlhoch.** (DOC)

**S3 File. PRISMA ScR Checklist** (DOC)

