## Supplemental Table 1 for "Transmission of SARS-CoV-2 on aircraft: A scoping review"

### Supplementary Appendix 1 – Search strategy for the assessment of flight-associated transmission of SARS-CoV-2

Ovid MEDLINE(R) ALL <1946 to November 02, 2023>

1	exp Coronavirus/	175240

2	exp Coronavirus disease 2019/	244781

3	exp Coronavirus infections/	256444

4	(Coronavir* or nCov or covid).mp.	404266

5	(("2019" adj (novel or new) adj corona*) or ("2019" adj (CoV or nCoV)) or (coronavirus adj (disease adj "2019")) or COVID19 or COVID-19 or ((Novel or New) adj Corona*) or SARS2 or SARS-CoV-2 or (SARS adj2 (coronaviridae or coronavirus)) or ((sars or Coronavirus) adj "2") or nCov or 2019ncov).mp.	393245

6	Coronaviridae Infections/ or Coronaviridae/ or SARS-CoV-2/ or COVID-19/	252431

7	((severe adj acute adj respiratory adj syndrome) or SARs or Sars-cov or ((sars-associated or sars-related) adj (cov or coronavirus))).mp.	229394

8	Betacoronavirus 1/ or Betacoronavirus/	33324

9	or/1-8	419890

10	exp Aircraft/	13149

11	exp Aerospace Medicine/	15178

12	exp Air Travel/	547

13	(aircraft or airplane* or aeroplane* or cabin* or flight or in?flight).mp.	102205

14	((air adj3 travel) or (flight adj5 passenger) or flying).mp.	12430

15	or/10-14	119668

16	9 and 15	1389

17	limit 16 to yr="2020-Current"	1248

Embase <1974 to 2023 November 02>

1	exp Coronavirus/	127315

2	exp Coronavirus disease 2019/	359672

3	exp Coronavirus infections/	379916

4	(Coronavir* or nCov or covid).mp.	482483

5	(("2019" adj (novel or new) adj corona*) or ("2019" adj (CoV or nCoV)) or (coronavirus adj (disease adj "2019")) or COVID19 or COVID-19 or ((Novel or New) adj Corona*) or SARS2 or SARS-CoV-2 or (SARS adj2 (coronaviridae or coronavirus)) or ((sars or Coronavirus) adj "2") or nCov or 2019ncov).mp.	468202

6	Coronaviridae Infections/ or Coronaviridae/ or SARS-CoV-2/ or COVID-19/	155454

7	((severe adj acute adj respiratory adj syndrome) or SARs or Sars-cov or ((sars-associated or sars-related) adj (cov or coronavirus))).mp.	207182

8	Betacoronavirus 1/ or Betacoronavirus/	7420

9	or/1-8	500807

10	exp Aircraft/	11651

11	exp Aerospace Medicine/	7880

12	exp Air Travel/	48345

13	(aircraft or airplane* or aeroplane* or cabin* or flight or in?flight).mp.	155334

14	((air adj3 travel) or (flight adj5 passenger) or flying).mp.	16015

15	or/10-14	178839

16	9 and 15	2587

Search Name:

Date Run:	03/11/2023 10:37:39

Comment:

ID	Search	Hits

#1	MeSH descriptor: [COVID-19] explode all trees	4894

#2	MeSH descriptor: [Severe Acute Respiratory Syndrome] explode all trees	389

#3	MeSH descriptor: [Severe acute respiratory syndrome-related coronavirus] explode all trees	2429

#4	(sars-associated OR sars-related OR Coronavir* OR nCov OR covid OR covid-19 OR Coronavirus OC43 OR HKU1 OR HCV-OC43 OR corona*)	89542

#5	#1 OR #2 OR #3 OR #4	89564

#6	MeSH descriptor: [Aircraft] explode all trees	187

#7	MeSH descriptor: [Air Travel] explode all trees	12

#8	(aircraft or airplane* or aeroplane* or cabin* or flight or in?flight or (air adj3 travel) or (flight adj5 passenger) or flying).mp.	20068

#9	#6 or #7 or #8	20264

#10	#5 AND #9 with Cochrane Library publication date Between Jan 2020 and Nov 2023, in Trials	291
