## Supplemental Table 2 for "Transmission of SARS-CoV-2 on aircraft: A scoping review"

### Supplementary Appendix 2 – Results of the Risk of Bias assessment of included studies related to flight associated transmission of SARS-CoV-2 as per the quality appraisal tool by Leitmeyer & Adlhoch^[1]^.

| **Author/Year** | **Index case classification** | **Secondary case ascertainment** | **Contact tracing strategy** | **Timeliness of contact tracing** | **Completeness of contact tracing** | **Limitations** | **Overall Score** | **Overall quality** |
| --- | --- | --- | --- | --- | --- | --- | --- | --- |
| **Dhanasekaran et al., 2021** | 1 | 2 | 2 | 2 | 2 | -1 | 8 | High quality |
| **Zhang et al., 2021** | 1 | 1 | 2 | 2 | 2 | 0 | 8 | High quality |
| **Bae et al., 2020** | 1 | 2 | 2 | 2 | 2 | -1 | 8 | High quality |
| **Murphy et al., 2020** | 1 | 2 | 0 | 2 | 1 | 0 | 6 | Medium quality |
| **Quach et al., 2021** | 1 | 2 | 0 | 2 | 2 | 0 | 7 | High quality |
| **Eldin et al., 2020** | 1 | 2 | 0 | 2 | 0 | -1 | 4 | Medium quality |
| **Blomquist et al., 2021** | 1 | 2 | 0 | 2 | 0 | 0 | 5 | Medium quality |
| **Yang et al., 2020** | 1 | 2 | 2 | 2 | 2 | -1 | 8 | High quality |
| **Zhang et al., XA 2020** | 1 | 2 | 0 | 2 | 0 | -1 | 4 | Medium quality |
| **Zhang et al., XA 2020** | 1 | 2 | 2 | 2 | 2 | -1 | 8 | High quality |
| **Butt et al., 2021** | 1 | 2 | 2 | 2 | 2 | -1 | 8 | High quality |
| **Schwartz et al., 2020** | 1 | 2 | 0 | 1 | 0 | 0 | 4 | Medium quality |
| **Hu et al., 2021** | 1 | 1 | 0 | 1 | 2 | -1 | 4 | Medium quality |
| **Nir Paz et al., 202** | 1 | 2 | 0 | 2 | 2 | -1 | 6 | Medium quality |
| **Swadi et al., 2021** | 1 | 2 | 0 | 2 | 2 | -1 | 6 | Medium quality |
| **Kong et al., 2021** | 1 | 2 | 0 | 2 | 2 | -1 | 6 | Medium quality |
| **Hoehl et al., 2020** | 1 | 2 | 0 | 0 | 0 | 0 | 3 | Low quality |
| **Choi et al., 2020** | 1 | 2 | 0 | 2 | 0 | -1 | 4 | Medium quality |
| **Eichler et al., 2021** | 1 | 2 | 0 | 2 | 0 | -1 | 4 | Medium quality |
| **Eichler et al., 2021** | 1 | 2 | 0 | 2 | 0 | -1 | 4 | Medium quality |
| **Pavli et al., 2020** | 1 | 2 | 2 | 1 | 2 | -1 | 7 | High quality |
| **Chen et al., 2020** | 1 | 2 | 2 | 2 | 2 | -1 | 8 | High quality |
| **Speake et al., 2020** | 1 | 2 | 0 | 2 | 0 | 0 | 5 | Medium quality |
| **Toyokawa et al., 2021** | 1 | 2 | 0 | 2 | 2 | -1 | 6 | Medium quality |
| **Lv et al., 2021** | 1 | 2 | 2 | 2 | 2 | -1 | 8 | High quality |
| **White et al., 2022** | 1 | 2 | 0 | 1 | 0 | -1 | 3 | Low quality |
| **Draper et al., 2020** | 1 | 2 | 0 | 0 | 1 | -1 | 3 | Low quality |
| **Park et al., 2020** | 1 | 2 | 2 | 2 | 0 | -1 | 6 | Medium quality |
| **Moek et al., 2022** | 1 | 1 | 0 | 1 | 0 | 0 | 3 | Low quality |
| **Khahn et al.,** | 1 | 2 | 2 | 2 | 2 | 0 | 9 | High quality |
| **Guo et al., 2022** | 1 | 2 | 2 | 2 | 2 | 0 | 9 | High quality |
| **Williamson et al., 2022** | 1 | 0 | 2 | 2 | 2 | -1 | 6 | Medium quality |
| **Ngeh et al.,** | 1 | 2 | 2 | 2 | 2 | 0 | 9 | High quality |

| **Scoring Criteria as per Leitmeyer & Adlhoch** | **Points Awarded or Withdrawn** |
| --- | --- |
| **Index case classification** | |
| Laboratory confirmation | 1 |
| Unspecific clinical diagnosis (based on symptoms) or not reported | 0 |
| **Secondary case ascertainment** | |
| Laboratory confirmation of all cases | 2 |
| Unspecific clinical diagnosis (based on symptoms) or no comprehensive confirmation of all secondary cases | 1 |
| Not provided | 0 |
| **Contact tracing strategy** | |
| Comprehensive (all passengers and crew) | 2 |
| Other (close contacts, highly exposed contacts etc) | 0 |
| **Timeliness of contact tracing** | |
| Within 1 week | 2 |
| Within 3 weeks | 1 |
| After 3 weeks or more | 0 |
| **Completeness of contact tracing: proportion of passengers followed up** | |
| More than 80% were followed up | 2 |
| Between 80% and 50% were followed up | 1 |
| Less than 50% were followed up or retrospective identification | 0 |
| **Limitations** | |
| Alternative exposure not addressed | -1 |

1. Leitmeyer K, Adlhoch C. Review Article: Influenza Transmission on Aircraft: A Systematic Literature Review. Epidemiology (Cambridge, Mass). 2016;27(5):743-51. Epub 2016/06/03. doi: 10.1097/ede.0000000000000438.
